# Association of ADAM33 Gene rs2280091 (T1) Polymorphism with Asthma Severity in Syrian Population: A Case-Control Study Using PCR-RFLP Analysis

**DOI:** 10.1101/2025.07.17.25331673

**Authors:** Joseph George Shenekji, Ghenwa Lbabidi, Abdullah Khoury

## Abstract

**Background:** Asthma is a complex chronic inflammatory disease affecting approximately 339 million people worldwide. The ADAM33 gene, encoding a disintegrin and metalloproteinase, has emerged as a key susceptibility gene for asthma, with the rs2280091 (T1) polymorphism showing variable associations across different populations. This study represents the first genetic investigation of asthma in the Syrian population.

**Methods:** A case-control study was conducted at Aleppo University Hospital from April to November 2019, including 100 participants (80 asthma patients and 20 healthy controls) aged 20-40 years. Asthma diagnosis was confirmed using spirometry and reversibility testing according to GINA guidelines. Genomic DNA was extracted from whole blood, and the rs2280091 polymorphism was genotyped using PCR-RFLP with NcoI restriction enzyme. Statistical analysis was performed using SPSS 25.0 with significance set at p≤0.05.

**Results:** The study population showed balanced sex distribution (50% male, 50% female) with mean ages of 26.13 years (cases) and 29.65 years (controls). Genotype frequencies were: A/A (43.0%), A/G (45.0%), and G/G (12.0%), with allele frequencies of A=0.66 and G=0.34, conforming to Hardy-Weinberg equilibrium. While no significant association was found between genotype and asthma occurrence (p=0.871), the G/G genotype showed significant association with increased asthma severity (p=0.016). ANOVA analysis revealed significantly lower FEV1 values in G/G carriers compared to A/A and A/G genotypes (p=0.001).

**Conclusions:** The ADAM33 rs2280091 G/G genotype is significantly associated with increased asthma severity in the Syrian population, suggesting its potential utility as a genetic marker for severe asthma phenotypes. This finding contributes to understanding asthma genetics in Middle Eastern populations and supports the role of ADAM33 in airway remodeling processes.

## 1. Introduction

Asthma is a complex chronic inflammatory disease of the airways characterized by variable and recurring symptoms, bronchial hyperresponsiveness, and reversible airflow obstruction. According to the World Health Organization, approximately 339 million people worldwide were affected by asthma in 2016, with 918,417 deaths attributed to the disease globally, representing 24.8 million disability-adjusted life years (DALYs) [22]. The burden of asthma-related mortality is disproportionately higher in low- and middle-income countries, highlighting the need for population-specific research to understand disease mechanisms and develop targeted therapeutic approaches.

The pathogenesis of asthma involves complex interactions between genetic predisposition and environmental factors. More than 127 genes have been identified as potentially associated with asthma susceptibility, distributed across 18 chromosomes, with many more awaiting discovery. These genes vary in their expression patterns and symptom severity associations across different ethnic groups and environmental conditions. Key asthma-associated genes include ADAM33, GSTM1, GSTT1, TNF, GSTP1, IL1RL1, SMAD3, RORA, ORMDL3, DPP10, TSLP, and IL13, each contributing to different aspects of asthma pathophysiology.

Among these genes, ADAM33 (A Disintegrin And Metalloproteinase 33) has emerged as one of the most extensively studied and significant asthma susceptibility genes [3]. Located on chromosome 20p13, ADAM33 was first identified through positional cloning in a landmark study by Van Eerdewegh et al. (2002) [3], which demonstrated its association with bronchial hyperresponsiveness in American and British populations. The gene encodes a zinc-dependent metalloproteinase that belongs to the disintegrin and metalloproteinase family, characterized by its role in cell-cell and cell-matrix interactions.

The ADAM33 protein contains several functional domains including a prodomain, metalloproteinase domain, cysteine-rich domain, epidermal growth factor-like domain, transmembrane domain, and cytoplasmic domain. These domains facilitate its involvement in various cellular functions, particularly in mesenchymal cells including airway fibroblasts; myoblasts, and smooth muscle cells. The protein’s unique combination of adhesive and proteolytic domains distinguishes it from other membrane proteins and positions it as a key mediator in airway remodeling processes [4].

The ADAM33 gene contains 22 exons and has been subject to extensive polymorphism studies across diverse populations. However, no single polymorphism has shown universal association with asthma across all ethnic groups, with variations observed based on geographic and ethnic genetic pools. Environmental factors also play a crucial role in modulating the relationship between ADAM33 polymorphisms and asthma phenotypes through gene-environment interactions and epigenetic mechanisms [29].

The rs2280091 (T1) polymorphism, located in the cytoplasmic domain of ADAM33, has been particularly well-studied. A comprehensive meta-analysis by Li et al. (2019) [2] examining 63 articles from 20 countries found variable associations between this polymorphism and asthma risk. The T1 polymorphism was studied in 34 case-control studies encompassing 7,873 asthma patients and 8,537 healthy controls, revealing population-specific differences in association patterns. A graphical summary is shown in figure 1.

**Figure 1.**
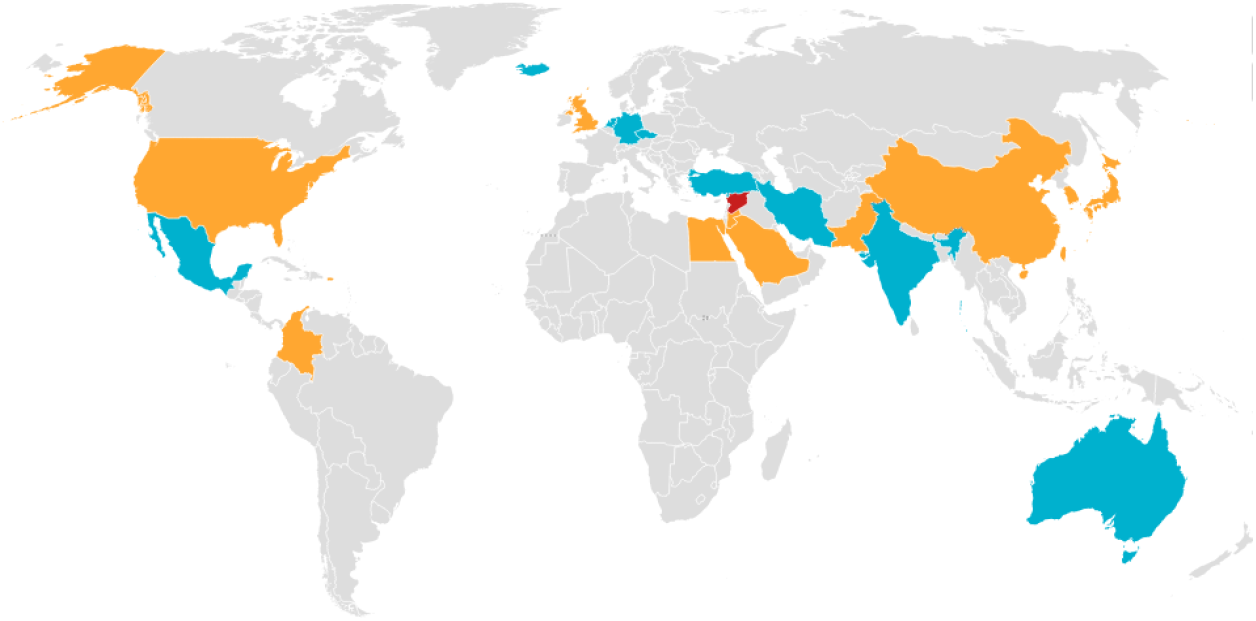
Global distribution of ADAM33 T1 polymorphism association studies showing countries with positive, negative, and no association with asthma. The light blue represents countries where there is no association of T1 with asthma, and the orange color represents countries where a positive association between T1 and asthma was found.

Previous studies have shown that certain ADAM33 polymorphisms, including T1, are associated with decreased FEV1 levels in adult asthma patients, suggesting a role in determining disease severity rather than just susceptibility [5]. The proposed mechanism involves ADAM33’s influence on airway remodeling through several pathways: (1) damaged epithelial cells secrete TGF-β1 in response to allergens, leading to mesenchymal stem cell hyperresponsiveness and abnormal airway remodeling; (2) active TGF-β1 directly modulates ADAM33 expression; (3) chemical irritants and allergens disrupt airway epithelium, activating innate immune cells through C-type lectin receptors, toll-like receptors, and aryl hydrocarbon receptors, leading to Th2 cell formation and increased ADAM33 mRNA expression; and (4) environmental modifications of genetic and epigenetic factors cause aberrant ADAM33 expression in epithelial cells, fibroblasts, and smooth muscle cells [28].

The functional significance of ADAM33 polymorphisms extends beyond simple disease susceptibility. Studies have demonstrated that IL-4 and IL-13 cytokines regulate ADAM33 mRNA expression by binding to the 3’ untranslated region in fibroblasts, with maximal activation occurring when both cytokines act together. Conversely, TGF-β2 downregulates ADAM33 mRNA expression through histone modification of the promoter region without affecting DNA methylation. These findings suggest that ADAM33 expression is tightly regulated by inflammatory mediators characteristic of asthmatic airways.

Population-specific studies have revealed interesting patterns in ADAM33 T1 associations. In Asian populations, including studies from China [11], Korea, Japan [13], and Taiwan [15], the T1 polymorphism has generally shown positive associations with asthma risk and severity. However, studies from European populations [14] have yielded mixed results, with some showing no association or even protective effects. Middle Eastern populations, including studies from Saudi Arabia [8], Egypt [9], and Iran [10], have shown predominantly positive associations, though with some variability.

The Syrian population represents a unique genetic background in the Middle East, with historical influences from diverse ethnic groups and limited previous genetic studies of asthma. Understanding the genetic architecture of asthma in this population is crucial for developing personalized therapeutic approaches and contributing to the global understanding of asthma genetics. This study represents the first investigation of ADAM33 polymorphisms in the Syrian population, focusing specifically on the rs2280091 (T1) variant and its association with asthma severity rather than just disease occurrence.

## 2. Methods

### 2.1. Study Design and Population

This case-control study was conducted at Aleppo University Hospital, Department of Internal Medicine, Pulmonary Function Laboratory, from April 9, 2019, to November 30, 2019. The study protocol was approved ethically and scientifically by the directorate of post-grad and scientific research in the University of Aleppo, syria (2018-2019) and all participants provided written informed consent before enrollment.

A total of 100 participants were recruited, including 80 asthma patients and 20 healthy controls, all aged 20-40 years and representing both Aleppo city and surrounding rural areas. The study population consisted of 50 males and 50 females to ensure balanced sex distribution. All participants were non-smokers to eliminate confounding effects of tobacco exposure on lung function and genetic associations.

### 2.2. Inclusion and Exclusion Criteria

Inclusion criteria for asthma patients included: (1) age 20-40 years; (2) non-smoking status; (3) confirmed asthma diagnosis by a specialist physician; (4) positive reversibility test demonstrating ≥10% improvement in FEV1 after bronchodilator administration; and (5) residence in Aleppo governorate. Exclusion criteria included: (1) negative asthma diagnosis; (2) chronic bronchitis or other chronic respiratory diseases; (3) active smoking or significant smoking history; (4) previous participation in the study; and (5) inability to perform adequate spirometry.

Healthy controls were selected from individuals visiting the hospital for routine check-ups or accompanying patients, with no history of asthma, allergic diseases, or chronic respiratory conditions. Controls underwent the same spirometry testing to confirm normal lung function.

### 2.3. Clinical Assessment and Spirometry

All participants underwent comprehensive clinical evaluation including detailed medical history, physical examination, and pulmonary function testing using a SPIROLAB III spirometer. Spirometry was performed according to American Thoracic Society/European Respiratory Society guidelines [24], with participants instructed to avoid meals for 2 hours, smoking for >1 hour, and bronchodilators for 6-24 hours before testing.

Asthma severity was classified according to Global Initiative for Asthma (GINA) guidelines [1] and American Academy of Family Physicians criteria based on FEV1 values: Mild (FEV1>70%), Moderate (FEV1 60-69%), Moderately Severe (FEV1 50-59%), Severe (FEV1 35-49%), and Very Severe (FEV1 <35%).

Reversibility testing was performed using salbutamol (200 μg) administered via metered-dose inhaler with spacer device. Post-bronchodilator spirometry was performed 15 minutes after drug administration. Reversibility was defined as ≥10% improvement in FEV1, based on European Respiratory Society criteria for 95% statistical confidence in excluding random bronchodilation.

### 2.4. Blood Sample Collection and DNA Extraction

Venous blood samples (4 mL) were collected in EDTA tubes to prevent coagulation. Samples were divided for complete blood count analysis (including eosinophil count determination using Sysmex analyzer) and genomic DNA extraction. DNA extraction was performed on the day of blood collection using the Vivantis GF-1 kit following manufacturer’s instructions.

The extraction protocol involved: (1) blood lysis using Buffer BB; (2) protein digestion with Proteinase K at 65°C for 10 minutes; (3) DNA precipitation with absolute ethanol; (4) column purification with sequential washes using Wash Buffer 1 and Wash Buffer 2; and (5) elution with pre-warmed elution buffer. DNA quality and concentration were assessed using 1% agarose gel electrophoresis and spectrophotometry (Eppendorf Biophotometer), with acceptable samples showing A260/A280 ratios between 1.8-2.0 and concentrations of 40-60 ng/μL.

### 2.5. PCR Amplification

PCR primers targeting the ADAM33 rs2280091 (T1) polymorphism were designed based on published sequences and validated using NCBI Primer-BLAST tool. The primer sequences were: Forward: 5’-ACTCAAGGTGACTGGGTGCT-3’ and Reverse: 5’-GAGGGCATGAGGCTCACTTG-3’, producing a 400 bp amplicon.

PCR reactions were performed in 25 μL volumes containing: 17.95 μL nuclease-free water,2.5 μL 10X PCR buffer, 0.75 μL MgCl2 (50 mM), 0.5 μL dNTPs (10 mM), 1 μL each primer (10 pmol), 0.3 μL Taq DNA polymerase (Genedirex), and template DNA. Thermal cycling was performed using Applied Biosystems thermal cycler with the following program: initial denaturation at 95°C for 5 minutes, followed by 37 cycles of 95°C for 30 seconds, 61°C for 30 seconds, and 72°C for 45 seconds, with final extension at 72°C for 7 minutes.

PCR products were verified by electrophoresis on 1.5% agarose gels in TAE buffer at 90V for 2 hours, using 50 bp DNA ladder as size marker (Genedirex). Products were visualized after ethidium bromide staining and UV transillumination. Shown in figure 2.

**Figure 2.**
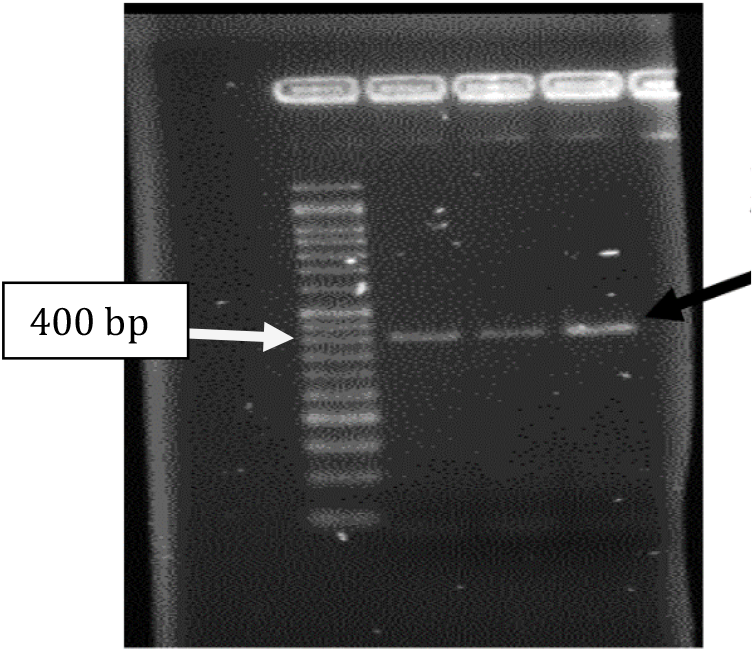
shows the PCR amplicon at 400 bp (pointed by the black arrow) that will be digested later to check for the polymorphism.

### 2.6. Genotyping by PCR-RFLP

The rs2280091 (T1) polymorphism was genotyped using restriction fragment length polymorphism (RFLP) analysis with NcoI restriction enzyme (Jena Bioscience). The enzyme recognizes the sequence 5’-CCATGG-3’ and produces different digestion patterns for each genotype.

Restriction digestion was performed in 20 μL reactions containing: 10 μL PCR product, 2 μL 10X restriction buffer, 0.5 μL NcoI enzyme (10 U/μL), and 7.5 μL nuclease-free water. Reactions were incubated at 37°C for 2.5 hours, followed by enzyme inactivation at 60°C for 20 minutes.

Digested products were analyzed on 3% agarose gels in TAE buffer at 90V for 2.5-3 hours. Genotypes were determined based on banding patterns: A/A genotype (wild-type) showed bands at 260 bp and 140 bp, G/G genotype (mutant) showed a single uncut band at 400 bp, and A/G genotype (heterozygous) showed all three bands (400 bp, 260 bp, and 140 bp). Shown in figure 3.

**Figure 3.**
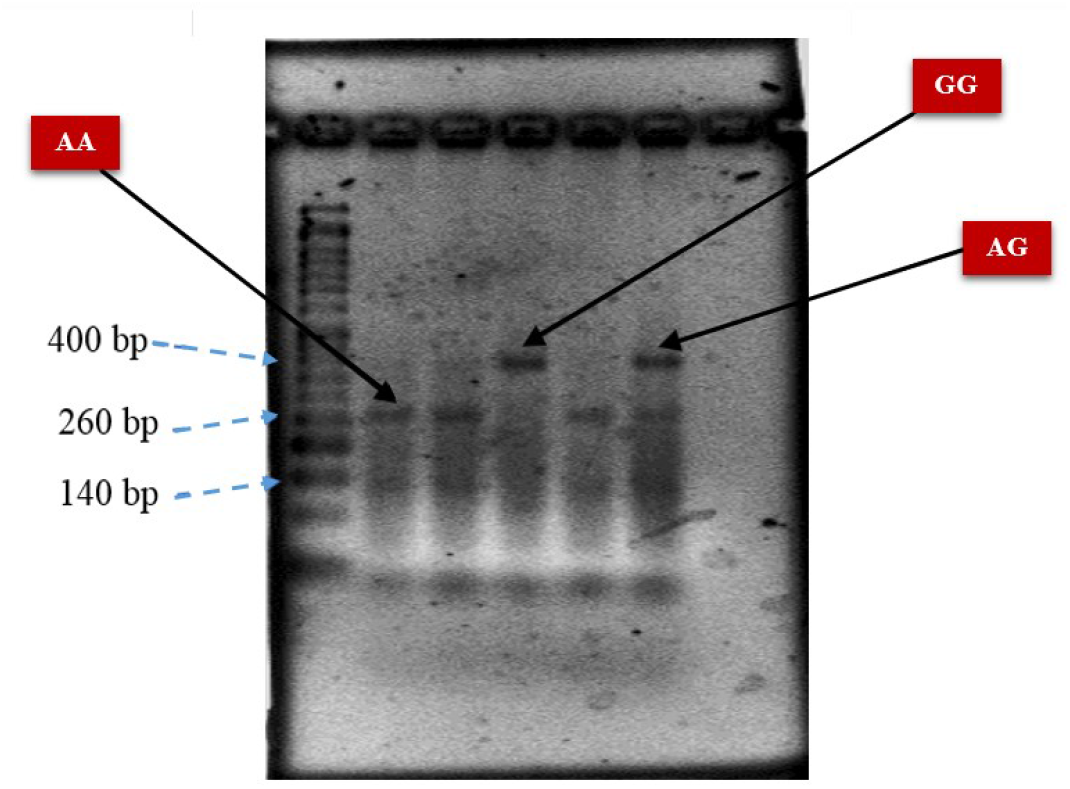
PCR-RFLP gel electrophoresis results showing genotype patterns for rs2280091 (T1) polymorphism.

### 2.7. Statistical Analysis

Statistical analysis was performed using SPSS version 25.0 software. Categorical variables were analyzed using cross-tabulation, Pearson’s chi-square test, and likelihood ratio (G2) statistics. Continuous variables were analyzed using one-way ANOVA with Tukey’s HSD post-hoc test for multiple comparisons. Hardy-Weinberg equilibrium was tested using chi-square goodness-of-fit test.

Odds ratios (OR) with 95% confidence intervals were calculated for genotype associations. Effect sizes were assessed using Cramer’s V and phi coefficients. Statistical significance was set at p ≤ 0.05. Allele and genotype frequencies were calculated using standard formulas, and Hardy-Weinberg equilibrium was tested using the equation: p^**2**^ + 2pq + q^**2**^ = 1.

Statistical significance was defined as p ≤ 0.05 with 95% confidence intervals. All tests were two-tailed.

## 3. Results

### 3.1. Demographic and Clinical Characteristics

The study included 100 participants with equal sex distribution (50 males, 50 females). The mean age was 26.13 years (SD ± 4.8) for asthma patients and 29.65 years (SD ± 6.2) for healthy controls, with no statistically significant difference between groups (p = 0.081). The age distribution showed the highest representation in the 19-23 years category (45%), followed by 24-28 years (28%).

Among asthma patients, 77% reported a family history of asthma, with equal distribution between maternal (35 cases) and paternal (33 cases) lineages, and 2 cases reporting asthma in both parents. This strong familial clustering was highly significant (p < 0.001) and suggests substantial genetic contribution to asthma susceptibility in the Syrian population [27].

The most common asthma triggers identified were fragrances and perfumes, psychological stress, and smoke exposure. Regarding occupational distribution, university students comprised the largest group (26%), followed by homemakers (19%), reflecting the demographic characteristics of the study area near university campuses.

### 3.2. Lung Function Parameters

Spirometry results showed significant differences between asthma patients and controls. The reversibility test demonstrated high diagnostic accuracy, with 87% of asthma patients showing ≥10% improvement in FEV1 after bronchodilator administration, compared to 5% of controls (p < 0.001). This finding validated the diagnostic criteria and confirmed the distinct physiological characteristics of the asthma group.

Asthma severity classification based on pre-bronchodilator FEV1 values revealed: 57.5% mild asthma (FEV1 >70%), 13.8% moderate asthma (FEV1 60-69%), 11.3% moderately severe asthma (FEV1 50-59%), 8.8% severe asthma (FEV1 35-49%), and 8.8% very severe asthma (FEV1 <35%). This distribution indicates that while most patients had mild disease, a substantial proportion (42.5%) had moderate to severe asthma.

### 3.3. Laboratory Parameters

Eosinophil count analysis revealed significant differences between asthma patients and controls (p = 0.004). Elevated eosinophil counts (>0.25 ×10^**3**^/μL) were exclusively observed in asthma patients, with 26% of patients showing eosinophilia compared to 0% of controls. This finding supports the allergic/atopic nature of asthma in many patients and provides additional validation of the case-control classification [26].

Weight analysis showed a significant association with asthma severity (p = 0.008), with patients in higher weight categories (>97 kg) showing representation across all severity levels, while patients with moderately severe asthma were prominently represented in the 80-96 kg weight category. This finding is consistent with previous studies linking obesity to asthma severity.

### 3.4. Genotype and Allele Frequencies

Successful genotyping was achieved for all 100 participants using PCR-RFLP analysis. The rs2280091 (T1) polymorphism showed the following genotype distribution: A/A (wild-type): 43.0% (n=43), A/G (heterozygous): 45.0% (n=45), and G/G (mutant): 12.0% (n=12). The allele frequencies were: A allele (major): 0.66 and G allele (minor): 0.34. as shown in table 1.

**Table 1.** Genotype and allele frequencies of rs2280091 (T1) polymorphism in study population.

| Phenotype frequency | Wild type allele | Heterozygous alleles | Mutant type allele |
| --- | --- | --- | --- |
|  | AA = 43 | AG = 45 | GG = 12 |
| Allele frequency | P (A) = 86 (0.66) |  | q (G) = 24 (0.344) |
| Genotype frequency | P<sup>2</sup> (AA)= 0.4356 | 2pq (AG)= 0.4554 | q<sup>2</sup> (GG)= 0.119025 |

Hardy-Weinberg equilibrium testing revealed that the observed genotype frequencies were in equilibrium with expected frequencies (χ^**2**^ = 0.0837, p > 0.01), indicating that the study population represents a stable genetic pool without significant population stratification or technical genotyping errors.

Comparison with previous studies revealed interesting population-specific differences. The G/G genotype frequency in our Syrian population (12.0%) was notably higher than reported in Chinese populations according to Shen et al., 2017 [11] but similar to some Middle Eastern populations, suggesting regional genetic similarities.

### 3.5. Association Analysis

#### 3.5.1 Genotype and Asthma Occurrence

Cross-tabulation analysis of genotype distribution between asthma patients and controls revealed no statistically significant association (G2 = 0.276, p = 0.871). The genotype frequencies in asthma patients were: A/A: 43.8% (35/80), A/G: 43.8% (35/80), and G/G: 12.5% (10/80). In controls: A/A: 40.0% (8/20), A/G: 50.0% (10/20), and G/G: 10.0% (2/20).

Similarly, analysis of G allele presence showed no significant association with asthma occurrence (G2 = 0.92, p = 0.761; OR = 1.167, 95% CI: 0.430-3.164). This finding suggests that while the rs2280091 polymorphism may not determine asthma susceptibility in the Syrian population, it may influence disease severity among those with asthma.

#### 3.5.2Genotype and Asthma Severity

The most significant finding of this study was the strong association between genotype and asthma severity. Cross-tabulation analysis revealed a highly significant association (G2 = 18.747, p = 0.016; phi = 0.452, p = 0.037). The G/G genotype showed a clear pattern of association with more severe asthma phenotypes. Displayed in figure 4.

**Figure 4.**
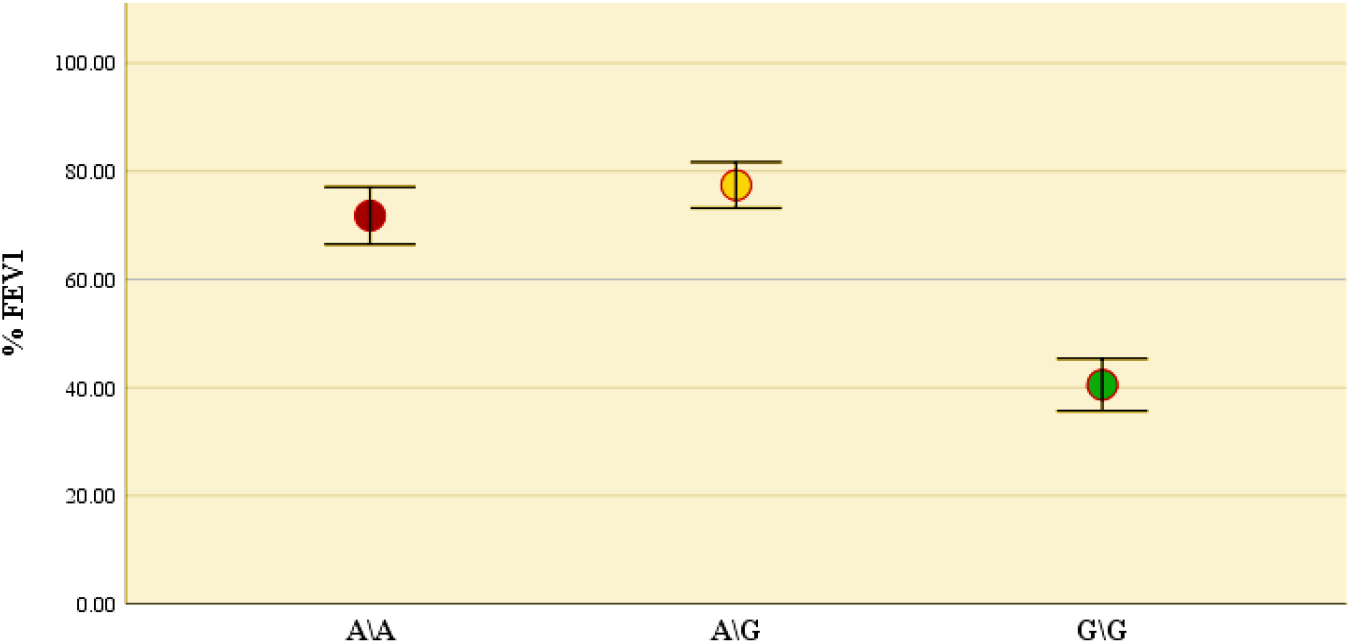
Simple Error Bar Plot to compare the mean asthma severity among carriers of the three genotypes for the studied T1 site, where G/G genotype shows increased asthma severity in comparison with other genotypes.

Analysis of FEV1 values by genotype using one-way ANOVA demonstrated significant differences between genotype groups (F = 8.309, p = 0.001). Post-hoc analysis using Tukey’s HSD revealed that G/G genotype carriers had significantly lower FEV1 values compared to both A/A carriers (mean difference = 16.048, p = 0.042) and A/G carriers (mean difference= 25.489, p < 0.001).

The mean FEV1 values by genotype were: G/G genotype: 52.00% (indicating severe asthma), A/A genotype: 68.05% (indicating moderate asthma), and A/G genotype: 77.49% (indicating mild asthma). This clear genotype-phenotype correlation supports the functional significance of the rs2280091 polymorphism in determining asthma severity. As mentioned in table 2.pe **N**

**Table 2.**
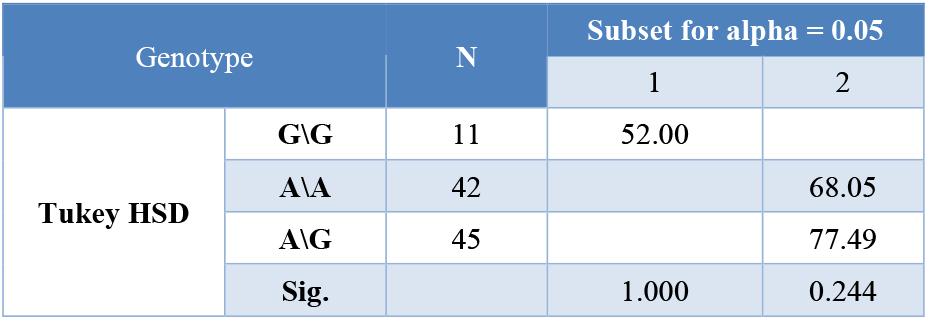
Association analysis between rs2280091 (T1) genotype and asthma severity parameter.

### 3.6. Additional Associations

Further analysis revealed that the rs2280091 polymorphism showed trends toward association with eosinophil count (G2 = 15.058, p = 0.058), approaching statistical significance and suggesting potential relationships with allergic inflammatory pathways. However, no significant associations were found with other demographic or clinical variables including age, sex, blood group, or occupational categories.

The lack of association between genotype and basic asthma occurrence, combined with the strong association with severity, suggests that the rs2280091 polymorphism may be more relevant to disease progression and phenotype determination rather than initial disease susceptibility.

## 4. Discussion

This study represents the first investigation of ADAM33 gene polymorphisms in the Syrian population and provides novel insights into the genetic architecture of asthma severity in Middle Eastern populations. Our findings demonstrate that while the rs2280091 (T1) polymorphism does not significantly influence asthma susceptibility, it shows a strong association with disease severity, particularly in individuals carrying the G/G genotype.

### 4.1. Genotype-Phenotype Correlations

The most important finding of our study is the clear genotype-phenotype correlation observed for asthma severity. Individuals carrying the G/G genotype demonstrated significantly lower FEV1 values (mean 52.00%) compared to A/A (68.05%) and A/G (77.49%) genotype carriers. This finding is consistent with previous studies from Iran by Karimi et al., 2014 [6] and the United States by Holgate et al., 2006 [5], which reported similar associations between homozygous G genotype and increased asthma severity.

The biological plausibility of this association is supported by the known functional consequences of the rs2280091 polymorphism. Located in the cytoplasmic domain of ADAM33, this polymorphism may alter intracellular signaling pathways, leading to abnormal fibroblast proliferation and smooth muscle cell hyperplasia—hallmarks of airway remodeling in asthma. The cytoplasmic domain of ADAM33 contains several potential phosphorylation sites that could be affected by this polymorphism, potentially altering protein-protein interactions and downstream signaling cascades.

Our findings contrast with some previous studies, particularly from Pakistan by Saba et al., 2018 [7], which reported a protective effect of the G/G genotype against asthma development. This discrepancy may reflect population-specific genetic backgrounds, different environmental exposures, or varying study methodologies. The Syrian population’s unique genetic history, influenced by multiple ethnic groups throughout history, may contribute to these distinct associations.

### 4.2. Population Genetics and Hardy-Weinberg Equilibrium

The observation that our study population was in Hardy-Weinberg equilibrium provides confidence in our genotyping results and suggests that the Syrian population represents a stable genetic pool without significant population stratification. The allele frequencies observed (A = 0.66, G = 0.34) are similar to those reported in other Middle Eastern populations, suggesting regional genetic similarities.

The relatively high frequency of the G/G genotype (12.0%) in our population, compared to East Asian populations (typically <5%), may contribute to the observed strong association with asthma severity. This higher frequency could reflect historical population admixture or founder effects specific to the Syrian population.

### 4.3. Clinical Implications

The strong association between G/G genotype and asthma severity has several clinical implications. First, genotyping for rs2280091 could potentially serve as a biomarker for identifying patients at risk of developing severe asthma, allowing for earlier intervention and more intensive monitoring. Second, understanding the genetic basis of asthma severity may inform personalized treatment approaches, as patients with different genotypes may respond differently to specific therapeutic interventions.

The finding that family history of asthma was present in 77% of our asthma patients, with equal contribution from maternal and paternal lineages, reinforces the importance of genetic factors in asthma development in the Syrian population. This high heritability suggests that genetic testing could be valuable for family counseling and risk assessment.

### 4.4. Mechanistic Considerations

The mechanism by which the rs2280091 polymorphism influences asthma severity likely involves multiple pathways. ADAM33 is known to be regulated by Th2 cytokines, particularly IL-4 and IL-13, which are elevated in asthmatic airways. The polymorphism may alter the responsiveness of ADAM33 to these regulatory signals, leading to dysregulated protein expression and function. [31]

Furthermore, ADAM33 expression is suppressed by TGF-β2 through histone modification mechanisms. The rs2280091 polymorphism might interfere with these epigenetic regulatory mechanisms, resulting in persistent ADAM33 expression and ongoing airway remodeling. This could explain why G/G genotype carriers show more severe disease phenotypes. [32]

The proteolytic activity of ADAM33 affects extracellular matrix turnover and cell migration, processes that are crucial in airway remodeling. The rs2280091 polymorphism may alter the balance between matrix synthesis and degradation, leading to excessive collagen deposition and smooth muscle proliferation characteristic of severe asthma. [33]

### 4.5. Environmental Interactions

The lack of association between genotype and asthma occurrence, combined with the strong association with severity, suggests that environmental factors may play a crucial role in determining which genetically susceptible individuals develop asthma. The Syrian population has experienced significant environmental changes in recent years, including increased air pollution, stress, and exposure to various allergens, which may interact with genetic predisposition to influence disease expression.

The finding that weight was significantly associated with asthma severity (p = 0.008) suggests that metabolic factors may interact with genetic predisposition to influence disease phenotype. Obesity is known to modify inflammatory pathways and may amplify the effects of genetic variants on asthma severity.

### 4.6. Limitations and Future Directions

Several limitations of this study should be acknowledged. First, the relatively small sample size (100 participants) may limit the statistical power to detect modest associations. Larger studies would be needed to confirm these findings and explore additional genetic variants and further confirm it. Second, the case-control design prevents assessment of temporal relationships between genotype and disease progression.

Future research should include longitudinal studies to track disease progression in relation to genotype, investigation of additional ADAM33 polymorphisms and haplotypes, and exploration of gene-environment interactions. Additionally, functional studies examining the biochemical consequences of the rs2280091 polymorphism would help elucidate the molecular mechanisms underlying the observed associations.

The development of larger, multi-center studies across different Syrian populations would help validate these findings and assess their generalizability. Investigation of other asthma-associated genes in the Syrian population would provide a more comprehensive understanding of the genetic architecture of asthma in this region.

## 5. Conclusion

This study provides the first evidence for the association between ADAM33 gene polymorphisms and asthma severity in the Syrian population. Our key findings include:

1. The rs2280091 (T1) polymorphism shows no significant association with asthma susceptibility but demonstrates a strong association with disease severity.
2. Individuals carrying the G/G genotype have significantly lower FEV1 values and more severe asthma phenotypes compared to A/A and A/G genotype carriers.
3. The Syrian population (Aleppo) shows distinct allele frequencies and genetic associations compared to other populations, highlighting the importance of population-specific genetic studies.
4. The strong familial clustering of asthma (77% positive family history) emphasizes the significant genetic component of asthma in the Syrian population.
5. PCR-RFLP analysis represents a cost-effective and reliable method for genotyping in resource-limited settings.

These findings contribute to our understanding of asthma genetics in Middle Eastern populations and suggest that the rs2280091 polymorphism could serve as a useful biomarker for identifying patients at risk of developing severe asthma. The results support the role of ADAM33 in airway remodeling processes and highlight the importance of genetic factors in determining asthma severity.

From a clinical perspective, these findings suggest that genetic testing for rs2280091 could be incorporated into asthma management protocols to identify high-risk patients who may benefit from more intensive monitoring and early intervention strategies. The strong association with severity rather than susceptibility indicates that this polymorphism may be particularly useful for prognostic purposes.

Future research should focus on replicating these findings in larger cohorts, investigating additional ADAM33 polymorphisms, and exploring the functional consequences of the rs2280091 variant. Understanding the complete genetic architecture of asthma in Syrian and other Middle Eastern populations will be crucial for developing personalized therapeutic approaches and improving patient outcomes.

## Acknowledgments

The authors thank the staff of Aleppo University Hospital, Department of Internal Medicine, for their assistance in patient recruitment and clinical assessments.

## Data Availability Statement

The datasets used and analyzed during the current study are available from the corresponding author upon reasonable request, subject to appropriate ethical approvals and data sharing agreements.

